# Whole Genome Sequencing Illuminates the Developmental Signatures of Human Language Ability

**DOI:** 10.1101/2021.11.22.21266703

**Authors:** Tanner Koomar, Lucas G Casten, Taylor R Thomas, Jin-Young Koh, Dabney Hofamman, Savantha Thenuwara, Allison Momany, Marlea O’Brien, Jeffrey Murray, J Bruce Tomblin, Jacob J Michaelson

**Affiliations:** Department of Psychiatry, University of Iowa; Molecular Otolaryngology and Renal Research Laboratories, University of Iowa; Biomedical Science Department, Iowa State University; Department of Psychological and Brain Sciences, University of Iowa; Department of Pediatrics, University of Iowa; Department of Communication Science and Disorders, University of Iowa

**Author notes:** https://www.overleaf.com/project/5e0663d86dd7730001dc3cfc.

## Abstract

Language is the foundation of human social interaction, education, commerce, and mental health. The heritability underlying language is well-established, but our understanding of its genetic basis — and how it compares to that of more general cognitive functioning— remains unclear. To illuminate the language-specific contributions of rare and common variation, we performed whole genome sequencing in N=350 individuals who were characterized with seven latent language phenotypes. We conducted region, gene, and gene set-based analyses to identify patterns of genetic burden that disproportionately explained these language factors compared to nonverbal IQ. These analyses identified language-specific associations with *NDST4* and *GRIN2A*, with common variant replication of *NDST4* in an independent sample. Rare variant burden analyses revealed three distinct functional profiles of genes that make contributions to language: a prenatally-expressed profile with enrichment for chromatin modifiers and broad neuropsychiatric risk, a postnatal cortex-expressed profile with enrichment for ion channels and cognitive/neuropsychiatric associations, and a postnatal, subcortically-expressed profile with enrichment of cilium-related proteins. Compared to a profile strongly associated with nonverbal IQ, these language-related profiles showed less intolerance to damaging variation, suggesting that the selection patterns acting on language differ from patterns linked to intellectual disability. Furthermore, we found evidence that rare potential reversions to an ancestral state are associated with poorer overall specific language ability. The breadth of these variant, gene, and profile associations suggest that while human-specific selection patterns do contribute to language, these are distributed broadly across numerous key mechanisms and developmental periods, and not in one or a few “language genes”.

## 2 Introduction

Language disorders are a common developmental challenge faced by up to 8% of children [63]. Without effective intervention, these conditions can negatively impact life-long academic and economic prospects [7]. Compromised language abilities are also a hallmark of several neurodevelopmental and psychiatric conditions [34]: from a high comorbidity of ADHD and language impairments [45], to the core symptomatology of autism, to the thought disorder endemic to schizophrenia [35, 36].

Beyond these well-documented developmental impacts of language disorders, the broader study of language ability can provide powerful insight into human cognition, brain development, and evolution. Language represents a complex set of cognitive functions that are distributed throughout the brain [20]. Understanding how human language manifests and develops may illustrate how diverse brain functions and circuits are integrated to carry out complex functions and behavior. Advanced communication through language also distinguishes humans from other species of great apes, functioning as a tool which allows us to organize into large social groups and societies. Consequently, insights into the roots and evolution of language ability are insights into what it means to be human.

To approach these questions, it is necessary to examine the full breadth of language ability which humans are capable of, rather than a disease-only perspective that scarifies substantial nuance and dynamic range. Certainly, some individuals have the “gift of gab”, and likely benefit from genetic mechanisms that contrast those with disordered language. Rather than impose *a priori* theoretical constructs to define language, we have taken a data-driven approach by deriving factors from a wide range of language and performance IQ tasks in the EpiSLI cohort.

EpiSLI represents one of the largest attempts to systematically study childhood language development and its impacts to date [62]. An epidemiological study of language impairment, the project began by screening over 7,000 kindergarteners across the state of Iowa for language impairments, and then fully phenotyping nearly 2,000 with a range of language and cognitive assessments. Several hundred of these subjects were followed throughout their primary and secondary school years and deeply phenotype for a variety of language constructs using age-appropriate instruments at regular intervals. We have, for the first time, generated whole genome sequencing on 350 individuals from this cohort.

Genetics represents a powerful tool for understanding the biology underpinning behavioral and cognitive phenotypes, like language, though this is not a trivial undertaking. Language is a multifaceted trait, and complex traits typically have complicated genetic bases. This is evident in the large and diverse set of genes proposed to impact language ability, with little replication success thus far [11, 34] This ambiguity arises through a combination of small sample sizes and a focus on individual common variants, whose effects are individually weak and distributed throughout the genome. Modern implementations of polygenic scores can, however, leverage the power of well-studied traits to provide insights into pleiotropic patterns of under-studied ones. Quantifying the effects of rare genetic variation at the gene level also poses its own set of difficulties. In particular — in the absence of variant segregation in pedigrees (which is only applicable in cases of severe disorder) — individual rare variants must be meaningfully collapsed or summarized at a higher level (e.g. genes). New and developing methods are thus needed to adequately address this class of variation.

In light of these challenges and opportunities, the maturing field of language genetics is faced with a number of foundational questions. Can molecular and genetic information be used to parse language phenotypes into distinct dimensions? What are the functional characteristics of genes linked to language ability? How can neurobiology knowledge be leveraged to illuminate the developmental components of language? Are there evolutionary features which inform the links between genetic variation and language ability? The answers to these questions will provide critical insight into the biological basis of human language. This study investigates these questions utilizing a powerful combination of a deeply phenotyped cohort, and whole-genome sequencing.

## 3 Results

### 3.1 Factor Analysis

Seven factors derived here represent diverse and distinct (though not orthogonal) components of language development from kindergarten (g0) through 4th grade (g4). The components derived from this analysis can be seen in Figure 1A. Factor F1 is driven by scores from the *sentence imitation* section of the TOLD-2P in kindergarten and the analogous *sentence repetition* sub-score of the CELF-III. F1 therefore appears to fairly specifically represent sentence processing, which has been found to be a robust indicator of overall language ability [32]. Factor F2 — primarily loading onto *receptive vocabulary* from the PPVT and *listening to paragraphs* from the CELF-III — seems to reflect receptive language at both small and large scales. Factor F3 is the only factor loading specifically to performance IQ at both kindergarten and 2nd grade and uniquely reflects this construct.

**Figure 1:**
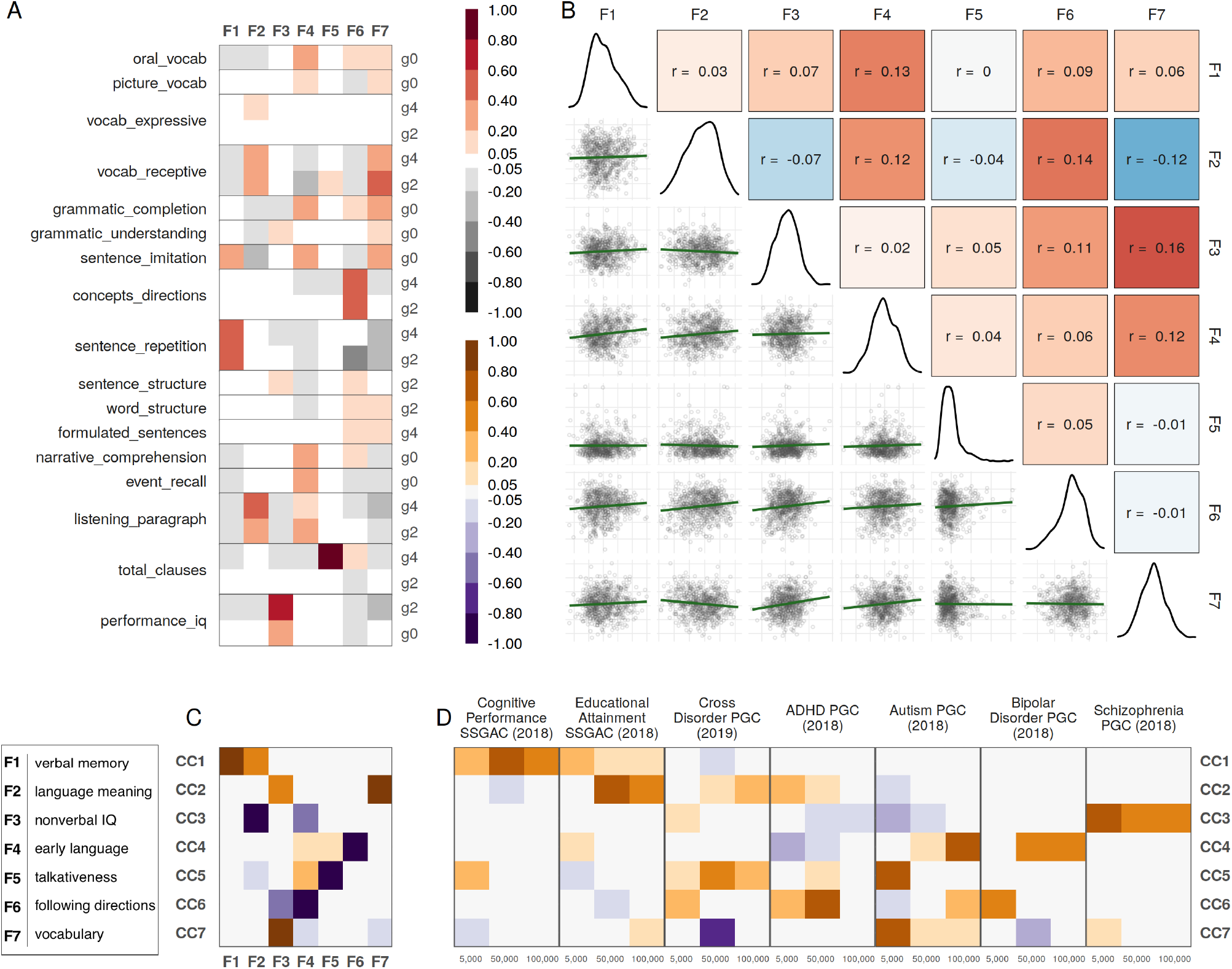
Language factors and polygenic score canonical correlation analysis. **A** Language factor components. **B** Pearson correlations for language factors (upper triangle), scatter plots of individual factor scores against one another (lower triangle), and distribution of each factor (diagonal). **C** CCA loadings on language factors. **D** CCA loadings on PGSes. Numbers on the x-axis indicate the approximate number of SNPs included in the PGS.

Factor F4 exhibits the strongest temporal association of all the factors, loading onto all kindergarten scores except for performance IQ. This may reflect early strength across all language domains, or it may result from the fact that most kindergarten assessments were derived from the TOLD-2P, while the assessments given at older ages were primarily derived from the PPVT and CELF-III. F4 is notably correlated with factors F1 and F2 (r = 0.13 and 0.12, respectively), but not F3 (IQ), suggesting that it is language-specific.

Factor F5 almost exclusively reflects the number of *clauses produced* in a narrative generation task [17], resulting in a heavily skewed distribution relative to other the factors (Figure 1B). While most students produced a number of clauses within a fairly narrow range, a small number produced several times more than most of their peers. Factor F5 has no notable correlations with other factors, possibly reflecting individual state when the assessment was taken.

Factor F6 is driven primarily by the *concepts and directions* assessment of the CELF-III, which appears to benefit from understanding of language meaning as well, as it is moderately correlated with F2 (r = 0.14). Factor F7 loads onto some of the broadest categories of assessments, from *receptive vocabulary* on the CELF-III and TOLD-2P to *grammar* and *word structure*. Notably, it has negative loadings on *sentence repetition* and *listening to paragraphs* (which primarily drive F1 and F2, respectively). F7 may therefore may represent command of the micro-structure of language, irrespective of working memory, or vocabulary more specifically.

### 3.2 Polygenic scores

The first canonical correlation component between the examined PGSes and language factors (CC1) loaded onto the overall language (F1) and language meaning (F2) factors (Figure 1C). CC1 loaded primarily onto PGS for cognitive performance (50,000 SNPs, Figure 1D), and to a lesser extent educational attainment (5,000 SNPs), illustrating that increasing genetic propensity for those traits are associated with better performance in overall language and interpretation of language meaning. In contrast, CC2 loaded primarily onto the performance IQ (F3) and vocabulary (F7) factors as well as PGS for educational attainment (50,000 SNPs), but *not* cognitive performance. CC2 also loaded somewhat robustly onto broad (cross-disorder) neuropsychiatric risk (100,000 SNPs) and ADHD (5,000 SNPs), possibly reflecting F7’s independence of working memory.

CC3 robustly loaded positively onto PGS for schizophrenia (5,000 SNPs) and negatively onto the early language (F4) and receptive language (F2) factors. This implies that genetic liability for schizophrenia is pleiotropic with generally compromised language ability, particularly receptive components, even at relatively young ages. Similarly, the loadings of CC4 suggests broad polygenic scores for bipolar disorder (100,000 SNPs) and autism (100,000 SNPs) is negatively associated with performance on the concepts and directions factor (F6), while CC5 suggests that higher-impact autism risk (5,000 SNPs), ADHD risk (50,000 SNPs), and broad (cross-disorder) neuropsychiatric risk (50,000 SNPs) are associated with reduced talkativeness (F5). CC6 loaded negatively onto both the IQ (F3) and early language (F4) factors and positively onto PGS for ADHD (50,000 SNPs), and to a lesser extent broad autism risk (100,000 SNPs) and higher-impact bipolar and cross disorder risk (5,000 SNPs). Finally, CC7 loaded positively onto IQ (F3) and autism (5,000 SNPs), and negatively onto broad (cross-disorder) neuropsychiatric risk (50,000 SNPs).

### 3.3 High-Impact Variants in Putative Language Genes

A total of 13 putative language genes were found to have rare, high-impact variants in individuals who scored low on at least one factor, and in no individuals who scores above average or high on those same factors (Table S5). The genes *ATXN10, AUTS2, CDH2, NECAB1, NFXL1, OPA3, PPP2R1B, WFDC1*, and *ZFYVE28* had high-impact rare variants in only low ability individuals, while the genes *DGKB, H2AFX, ROBO1*, and *SCN9A* had high-impact rare variants in at least one low-ability and one below average-ability individual. Across these genes, low scores on F7 (vocabulary) were the most represented (5 out of 13 genes), followed by F2 (receptive language) and F4 (early language) with 4 out of 13 genes. F5 (talkativeness) was the least represented factor. A low F5 score was found in a single individual (who also scored low on F1 and F4) with a high-impact variant in *NFXL1*. This may reflect the state-dependence of F5, or the fact that it primarily captures high performance, which was not a criteria for selection here. Details of all these variants can be found in Table S6.

### 3.4 Gene-level Common & Rare Variant Association

To assess the potential combined impacts of common and rare-variation on each of the 7 factors described above, we employed the Sequence Kernel Association Test (SKAT) [39] at the gene level. Unless otherwise noted, all tests were of the combined effects of common and rare variation, and FDR correction was applied across all factors and protein-coding genes (n = 124,464).

#### 3.4.1 Putative Language Genes

Of the putative language genes identified in [34], two were found to have FDR adjusted SKAT p-values less than 0.2 (Table S7). Variation in *GRIN2A* was associated with the talkativeness factor (F5, p = 0.00002, FDR = 0.065), while *NDST4* was associated with the early language factor (F4, p = 0.00003, FDR = 0.078). *NDST4* was identified as a candidate language gene via a small GWAS of language impairment [13], although it failed to replicate in an independent sample. *GRIN2A* has been linked to epileptic aphasia [4], and a rare *de novo* variant was observed in one individual with severe language impairment [5].

In the case of *NDST4*, subsequent investigation revealed common variation primarily contributed the SKAT signal (Figure S2). To assess the robustness of this loci, genotypes around *NDST4* were extracted from 111 European-ancestry individuals who were not included in the whole genome sequencing analysis, but for whom SNP array data was previously generated. After correcting for the first 20 genetic principal components, two SNPs in the region had nominally significant linear associations with F4: rs114239532 (NC_000004.11:g.115862342G>A, p = 0.0022, MAF = 0.018) and rs62317200 (NC_000004.11:g.115741885C>T, p = 0.033, MAF = 0.023).

#### 3.4.2 Protein Coding Genes

Among all protein coding genes, there were a number of suggestive (FDR < 0.2) associations between combined common and rare genetic variation and factor scores (Table S8). Factor 2 (receptive language) showed suggestive associations with a set of genes located on chromosome 1p35.2 including *SERINC2, SNRP40, ZCCHC17, FABP3*, and *NKAIN1* (FDR range = 0.017–0.18). Microduplication of 1p35.2 has been found in an ASD cohort, identifying SERINC2 as a candidate ASD gene [26]. Deletions of 1p35 are also associated with developmental delays and hearing impairment [68]. Expression of genes in the NKAIN1–SERINC2 region has been linked to modulation of neurotransmitter and metabolic pathways [72]. The *NKAIN1* protein is highly expressed in the hippocampus and the cerebellar granular cell layer of the brain [23]. Of note, the *FABP3* gene has been linked to schizophrenia and ASD, mice with a *FABP3* knockout displayed decreased social memory and novelty seeking [57].

F3 (IQ) had suggestive associations with the genes *MATN3* (p = 0.00004, FDR = 0.088) and *DRC1* (p = 0.00014, FDR = 0.197). *MATN3*’s function is primarily understood to involve formation of the extracellular matrix in a variety of tissues, and is expressed in the brain. MATN3 has been shown to directly interact with several signalling factors important for brain development via yeast two-hybrids [55], including NOTCH2NLA [19, 18].

In addition to the putative language gene *NDST4*, F4 (early language) had suggestive SKAT association with *ASTE1* (p = 0.00008, FDR = 0.144). *ASTE1* overlaps with — and is believed to impact the splicing and expression of — the gene *ATP2C1* [44], itself a paralogue of the putative language gene *ATP2C2* [59, 14]. Additionally, ASTE1 has been shown by affinity-purification mass spectrometry to interact with several proteins [27] which regulate axonal growth [70] and dendrite morphogenesis [21].

The highly skewed distribution of the F5 (talkativeness) lead to substantial p-value inflation in the SKAT test. This — combined with the potential state-dependence of the trait — means care must be taken not to over-interpret the large number of apparent associations with this factor. Notably, the strongest signal of F5 was with *GRIK4* (p = 0.000000069, FDR = 0.006), a glutamate-gated ionic channel (specifically, the KA1 kainate receptor subunit). Deletion of *GRIK4* has been linked to protection from bipolar disorder [33], while common variants in the region have been linked to ADHD [69] and major depression [52].

The sole suggestive association of F6 (concepts and directions) was with the poorly annotated *C19orf69* /*ERICH4* (p = 0.0001, FDR = 0.158). Meanwhile, F7 (vocabulary) had two suggestive associations with the immuoglobulin-like *LRGIN3* (p = 0.00002, FDR = 0.059) and mannosidase *MAN2B2* (p = 0.00012, FDR = 0.179)

### 3.5 Singleton Variants

To leverage the large number of unique (singleton) variants identified in our WGS data, we needed an estimate of a variant’s potential impact on each language factor. Because human language is a (relatively) recent phenomenon, we cannot assume that strong cross-species conservation is the primary hallmark of language-impacting variation, thus disqualifying most currently-available variant prioritization scores. Consequently, we opted to tune combinations of these metrics using machine learning and score each variant according to these optimized and generalizable criteria. Briefly, for each language factor, we trained a random forest classifier to identify whether singletons near putative language and cognitive genes belonged to individuals with low scores for that factor. The trained classifier was then applied to all singletons genome-wide (out-of-bag predictions were used for those singletons in the training set). The features used to produce these predictions represented a wide range of functional, pathogenic, and evolutionary genomic annotations.

#### 3.5.1 Feature/Annotation Importance

Depending on the language factor in question, singleton variants which were predicted to confer high levels of risk exhibited distinct patterns of enrichment for the genomic annotations on which the model was trained. Here, high-risk singletons are defined as the 33% that conferred the highest risk (for scoring poorly on the indicated factor), while low-risk variants are the 33% that conferred the lowest risk. These categories — as well as the middle 33% of variants — were further sub-divided into five bins for the purpose of visualization in Figure 3. All Z-statistics specifically reported here had a false discovery rate corrected p-value less than 0.05 and are based on out-of-bag predictions.

##### Variant Pathogenicity Scores

Measures of variant pathogenicity (CADD, FATHMM coding and FATHMM non-coding) were universally the most important features used by the random forest to produce final risk scores, though the particular pattern differed among factors. F1 was the only factor for which all three scores were substantially enriched among high risk alleles (max CADD Z = 12.7, max FATHMM noncoding Z = 36.1, max FATHMM coding Z = 13.4). F2 had inconsistent CADD enrichment in the highest risk variants, while both FATHMM scores were consistently enriched (max CADD Z = 11.2, max FATHMM noncoding Z = 20.9, max FATHMM coding Z = 10.8). Only the FATHMM noncoding score showed strong enrichment in IQ factor 3 (max FATHMM noncoding Z = 18.1), while for the early language F4, all three were strongly enriched among only the very highest-risk singletons (max CADD Z = 37.3, max FATHMM noncoding Z = 27.6, max FATHMM coding Z = 15.8). For F5 (talkativeness), moderately high risk-increasing singletons had elevated pathogenicity scores (max CADD Z = 8.6, max FATHMM noncoding Z = 5.6, max FATHMM coding Z = 10.1), while the very highest-risk variants were depleted for some scores (CADD Z = -9.3, FATHMM noncoding Z = -13.3). Singletons conferring the greatest risk for low F6 scores had elevated FATHMM coding (Z = 59.5) and CADD scores (Z = 37.1). F7 risk alleles had low-to no pathogenicity score enrichment for all but the highest risk variants, which were enriched for high CADD (max Z = 11.1) and FATHMM coding (Z = 17.8) scores, but lower FATHMM noncoding scores (Z = -8.2).

##### Coding Regions

Many of the most interesting gene-level impacts (e.g. frameshifts, premature stop codons) were too rare to test for enrichment, and had low importance in the model due to their sparsity. The gene-level impacts which were suitably common to be evaluated included flanking regions (upstream/downstream), introns, and common coding changes (missense/synonymous variants). Only Factors F3 and F5 demonstrated a pronounced enrichment of intronic singletons among the highest-risk conferring alleles (Z = 12.1 and Z = 13.6, respectively). Coding features — and to a lesser extent flanking regions — were more commonly enriched among high-risk singletons. In fact, except for F3, all factors had a significant enrichment of missense variants among high-risk alleles. Notably, the variants conferring the very greatest risk F4 and F7 were extremely enriched for missense variants (Z = 31.0 and Z = 32.7, respectively).

##### Conservation

PhastCons [66] is basepair-level measure of conservation based on the alignment of genomic sequence from 100 species of vertebrates with known phylogenetic relationships. This score is particularly enriched among high-risk singletons of F1 (max Z = 28.5), F4 (max Z = 23.1), and F6 (max Z = 43.9). For factors F2, F3, and F5, PhastCons had greater enrichment among moderate-to-low risk variants.

Comparing the singletons’ reference and alternate alleles to the reference chimpanzee allele can provide one level of evidence for potential human-specific function. Singletons where the alternate allele is the same as the chimpanzee allele (“Alt Chimp”) indicate potential reversion to the ancestral allele. By contrast, variants where the human and chimpanzee reference alleles are shared (“Ref Chimp”) indicates conservation as far back as the last common ancestor — though it does not preclude alleles derived early in the human-chimpanzee lineage.

Across each factor, three distinct patterns of enrichment for these alleles are apparent. Low risk singletons may be enriched for Ref Chimp (conserved) alleles, while high risk singletons are enriched for Alt Chimp (possible reversion) alleles. This pattern is seen in F1 and F7 (max Z for Alt Chimp = 14.4 at 93.3 percentile of risk, and Z = 26.1 at 86.6 percentile of risk, respectively). Low risk singletons may be enriched for Alt Chimp (possible reversion) alleles, while high risk singletons are enriched for Ref Chimp (conserved) alleles, as seen in F5 and F6 (max Z for Ref Chimp = 15.2 at 93.3 percentile of risk and Z = 15.7 at 80 percentile of risk, respectively). The most complex pattern is found in F2 and F3. For these factors, the very highest *and* lowest risk singletons are enriched for Ref Chimp (conserved) alleles, while Alt Chimp (possible reversion) alleles are of moderate-to-high risk for low ability. Factors F4 and F7 slightly resemble this pattern, but only for the singletons conferring the very highest amount of risk (approximately the top 5% in both cases).

A more nuanced quantification of conservation states across the genome are available in ConsHMM [2], which identifies 100 *de novo* conservation states across the human genome, based on multiple sequence alignment. A range of these conservation states are enriched for high risk variants, ranging from ancient (low ConsHMM number) to relatively recent (high ConsHMM number), as seen in Figure S3. Annotations of these states — the 15 with the most consistent enrichment among high-risk singletons for all factors — as reported by [2] are described in Table 0.1. Lower-numbered ConsHMM states generally over-represent protein coding and promoter sites, while higher-numbered sites are more likely to over-represent repeat regions and retrotransposons. For example, high-risk singletons for factors F1 and F3 are highly enriched for for ConsHMM state 76 (max Z = 61.7 and Z = 39.2, respectively), which contains more LTRs than any other ConsHMM state.

##### Gene-level Singleton Summary Score

To further describe the functional and developmental patterns associated with singleton risk, a gene-level singleton summary score was calculated based on the association between individual factor scores and individuals’ maximal singleton risk within each protein-coding gene (see methods for description of weighted least squares models). As expected, these statistics were more likely to be negative (*χ*^2^ = 309.1, p-value < 2e-16), indicating greater estimated singleton risk in most genes is associated with lower language (or cognitive ability in the case of F3). Individual gene-factor associations are given in Supplemental Table SX. Briefly, two genes were associated with F1 at *FDR <* 0.05 (*RIPK4, CLDN23*), none with F2, one with F3 (*NEUROD2*), two with F4 (*TECPR1, FAM27D1*), and none with F5, F6, or F7.

##### Derivation of Brain-relevant Gene Archetypes

Archetypal analysis was used to generate a set of 20 brain-relevant archetypes based on spatio-temopral brain expression, brain cell-type specific expression, and gene-level GWAS association.

Two of these archetypes (A1 and A2) represented genes which conferred broad neurodevelopmental risk via association with the gene-level singleton summary scores for multiple language and cognitive factors (Figure 2A). Genes scoring high for archetype A1 were highly expressed across the prenatal cortex and striatum, with elevated expression in all classes of tested neurons (Figure 2C,D). A2 described a similar pattern of gene expression, albeit to a lesser extent than A1 (Figure 2D). Expression in all non-neuronal cell types was strongly associated with A2, while A1 was limited to associations with astrocytes and OPCs (Figure 2C).

**Figure 2:**
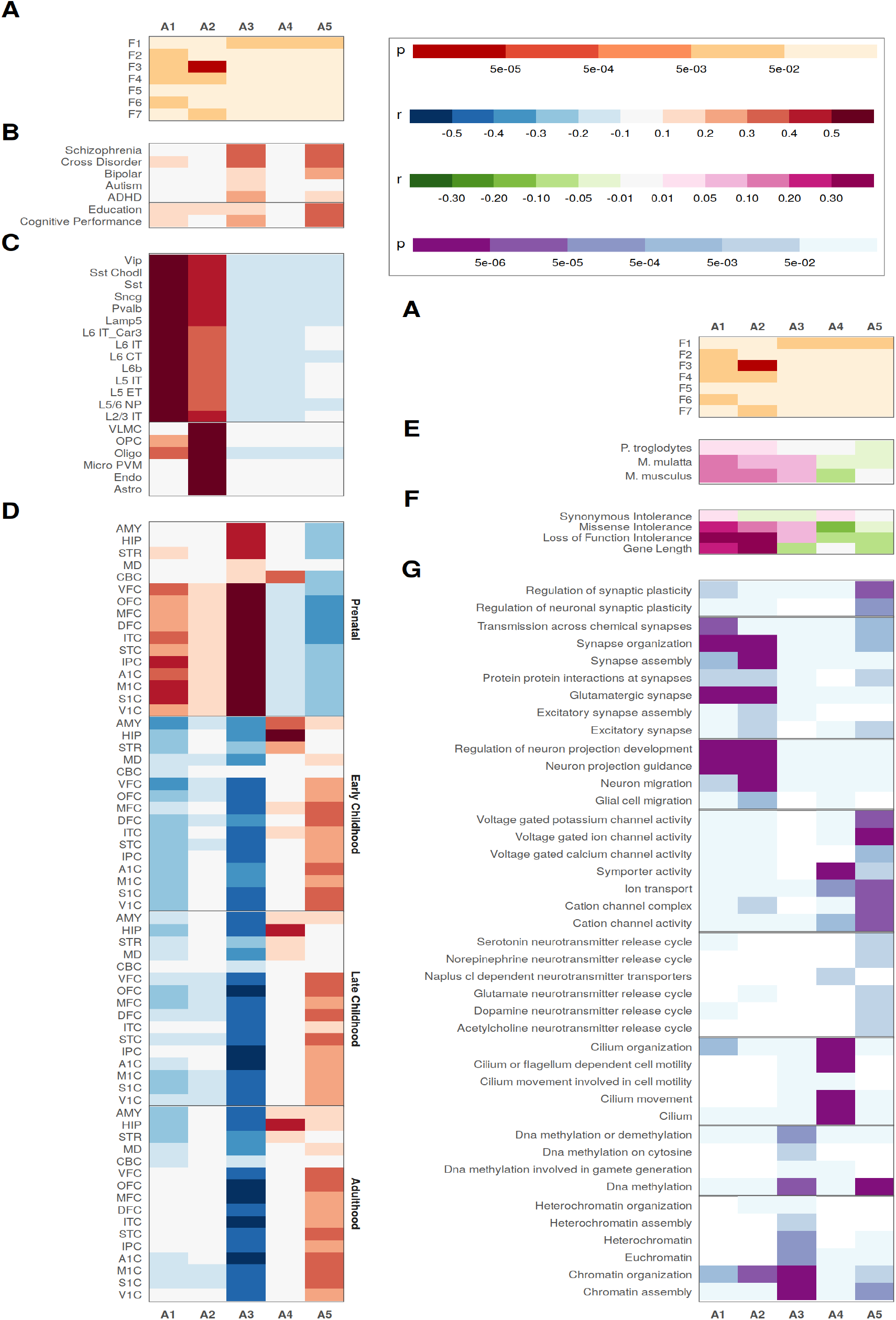
Archetypal description of gene-level singleton risk scores. **A** P-value from the one-sided Pearson correlation between the indicated archetype and the gene-level singleton risk score. **B** Pearson’s R between the indicated archetype and MAGMA results from GWAS for the trait indicated on the y-axis. **C** Pearson’s R between the indicated archetype and average cell-type specific gene expression. **D** Pearson’s R between the indicated archetype and average gene expression in the brain region and time-point based on the Brainspan developmental transcript. **E** Pearson’s R between the indicated archetype and sequence-level percent identify for the species indicated on the y-axis. **F** Pearson’s R between the indicated archetype and population-derived gene-level constraint metrics from gnomAD. **G** P-value from gene ontology and Reactome pathway gene set enrichment for the top 5% of genes for the indicated archetype.

Three other archetypes (A3–A5) displayed specificity for factor F1 (overall language) (Figure 2A). A3 described genes highly expressed across the entire prenatal brain, including sub-cortical regions (Figure 2D). A3 reflected GWAS signal for all included traits, particularly schizophrenia, psychiatric cross-disorder, cognitive performance and ADHD (Figure 2B). A4 was primarily defined by genes highly expressed in the postnatal midbrain, particularly the hippocampus (Figure 2D). Genes scoring high for A5 had elevated postnatal cortical expression and lower prenatal expression. A5 was also informed by GWAS results, particularly for schizophrenia, psychiatric cross-disorder, bipolar disorder, educational attainment, and cognitive performance (Figure 2B).

The patterns of the remaining archetypes can be found in Figure S4.

##### Gene Set Enrichments

The five archetypes described above were found to reflect distinct patterns of functional constraint, evolutionary divergence, and pathway enrichment (Figure 2E-G). The broad neurodevelopment archetypes scores (A1 and A2) were significantly correlated with genes’ intolerance to missense and loss of function mutations, as well a gene-length (Figure 2F). The archetypes specifically associated with Factor F1 singleton risk (A3–A5) had little to no relationship with functional constraint.

The archetypes defined by elevated prenatal brain expression (A1-A3) also reflected evolutionary conservation (Figure 2E). This was indicated by significant positive correlations with the genes’ percent identity with homologs from mouse and macaque, and to a lesser extent with chimpanzees. Archetypes A4 and A5 — both reflecting postnatal brain expression — displayed more sequence-level divergence by comparison (particularly A4).

The broad developmental archetypes A1 and A2 tagged genes linked to synapse assembly and organization, neuron projection and migration (Figure 2G). The F1-associated archetypes (overall language) were enriched for more terms related to neuronal plasticity and homeostasis including: synaptic plasticity (A5), cilium (A4), voltage-gated ion channels (A5), ion transport (A4 and A5), chromatin organization and transcriptional regulation (A3).

## 4 Discussion

This whole genome sequencing study of 350 deeply phenotyped children allowed us to parse language ability into seven distinct factors, identify associations with polygenic scores for cognitive traits and neuropsychiatric conditions, and associate rare variation with individual differences in distinct aspects of language at the gene level. Importantly, the results presented here suggest that the capacity for human language is not determined entirely by the pathways and mechanism underlying general cognition (nonverbal IQ). We also find substantial overlap in the neurobiological and genetic roots of these traits, suggesting the observed differences speak to the foundations of human uniqueness.

We decomposed the language and cognitive assessments into a total of seven factors. The interaction of these factors with polygenic scores for neurocognitive traits was mediated by seven canonical correlation components (CCs), highlighting nuanced relationships between genetic risk and performance on tasks that may span multiple language domains. Surprisingly, a correlation component (CC2) which loaded onto both IQ (F3) and vocabulary (F7) was highly associated with polygenic score for educational attainment (EA-PGS), but *not* cognitive performance (CP-PGS). In contrast, the component (CC1) representing broad language skills including overall language (F1) and receptive language (F2) — loaded more onto both, but more on CP-PGS than EA-PGS. This finding supports observations described in [22], which found EA-PGS to be substantially more effective than CP-PGS at predicting multiple measures of non-verbal IQ (matrix reasoning and numerical intelligence). Further supporting our observations, findings in [22] found CP-PGS was more predictive of verbal IQ scores than EA-PGS. Together, these results suggest that the genetic mechanisms shared between IQ and language ability may emerge more in sustained, long-term manifestations of cognitive ability than in-the-moment, objectively measured cognitive performance, which seems to relate more to language.

A number of distinct evolutionary profiles related to overall language (F1) and nonverbal IQ (F3) were also apparent based on the functional characteristics of genes implicated by rare-variant analysis. Rare variants (i.e., singletons) that were possible reversions to the ancestral alleles — where the human alternate allele was the chimpanzee reference allele — provided significant risk for poor overall language (Figure 3). High-risk variants for poor overall language also had elevated PhastCons scores — perhaps driven by the fact that they were more likely to impact coding sequence (missense variants), which have systematically higher PhastCons scores than noncoding regions. The pattern of these associations is even more intriguing for IQ, where possible reversions confer moderate risk, while more-conserved singletons have either no effect, or induce profound risk. This results in more modest associations of high-risk variants for IQ with PhastCons than for overall language, probably because these variants were more likely to be located in introns or gene-flanking regions. It is important to note that these observations primarily target human-chimp divergence, while much of the groundwork for human brain evolution was laid earlier in the primate lineage. Some evidence of this can be found in the substantial enrichemnt of ConsHMM state 76 among high-risk variants for both F1 and F3 (Figure S3). This state contains more LTRs than any other. Such retrotransposon regions are believed to have provided important genomic material for primate brain evolution [8, 47] and development [46, 28]. These trends paint a picture of language traits driven by more “active” (coding) changes deriving their impact from regions under recent selective pressures, while traits driven by regulatory disruptions may be older.

**Figure 3:**
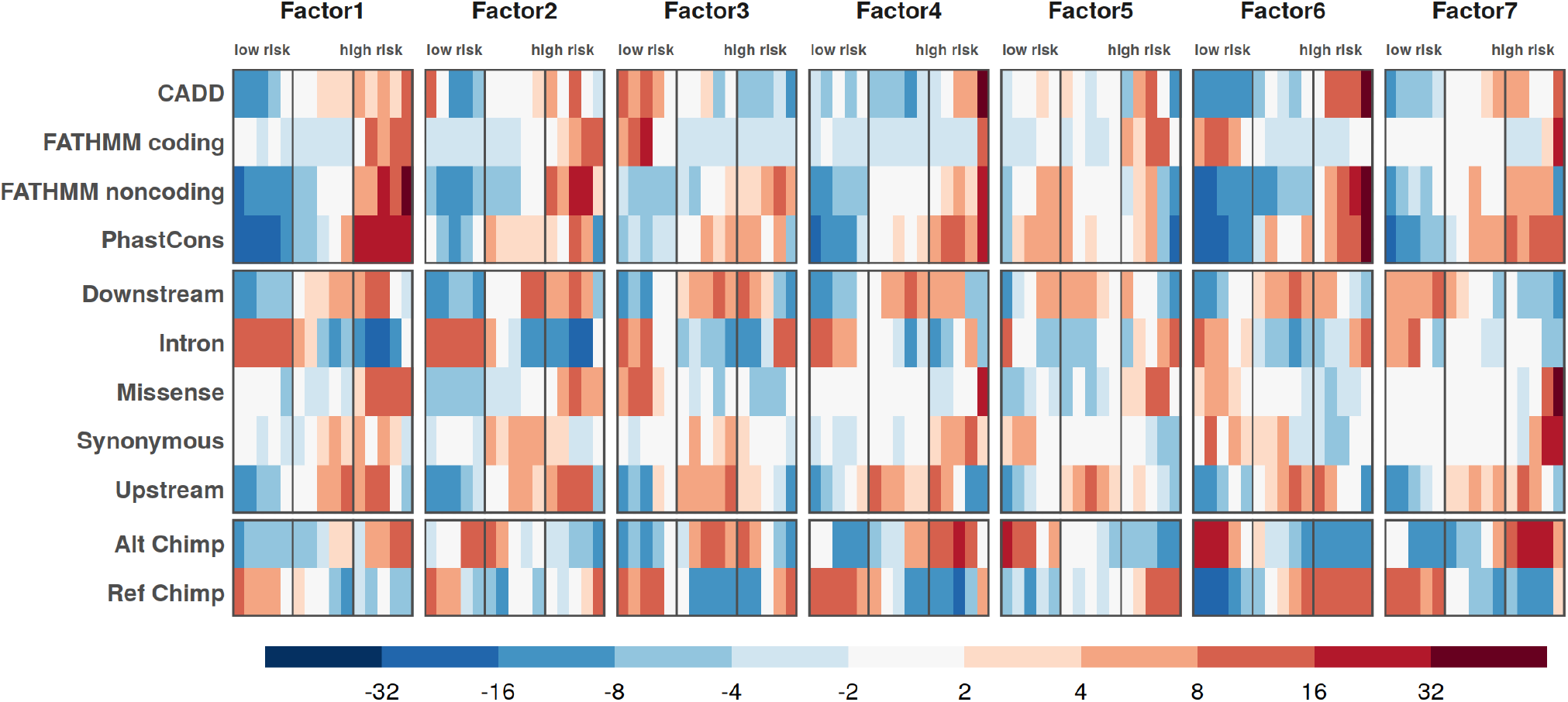
Association of selected variant annotations with increasing out-of-bag predictions of singleton risk, as quantified by logistic regression Z-statistic. ‘Ref Chimp’ indicates singletons where the reference allele is shared between humans and chimpanzees, while ‘Alt Chimp’ indicates sites where the alternate allele of the singleton is the chimpanzee reference allele.

Similar patterns were also apparent when comparing developmental and cell-type specific gene-expression profiles (archetypes) associated with rare-variant risk for broad neurodevelopment, compared to those for overall language alone. We found prenatal expression — particularly enriched in neurodevelopmental archetypes — was associated with greater evolutionary conservation and regulation of chromatin state. Postnatal expression patterns associated with overall language — especially high expression in the hippocampus — reflect greater sequence-level divergence and regulation of synaptic plasticity and neuron homeostasis. This again highlights that language abilities not only build upon on older patterns of early brain development, but arise from patterns of active brain maturation and maintenance.

Overall, this work provides clear support for the perspective of language ability as a multidimensional, polygenic trait impacted by both common and rare variation. We further find evidence of pleiotropic effects linking aspects of language with general cognitive ability and neuropsychiatric conditions, as evidenced from our analysis of polygenic scores. Complementary rare variant analysis likewise pointed to shared molecular and developmental profiles with more extensively studied neuropsychiatric and cognitive traits. Still, genes impacting language, and the characteristics of the associated genetic variants, point to more relaxed selective pressures compared to those acting on neurodevelopmental conditions like autism and intellectual disability. In order to adequately parse this genetic signal, larger sample sizes focusing on the full breadth of language ability (and not simply disorders) will be needed to make meaningful advances in distinguishing general neurodevelopmental vs. more language-specific effects. Finally, the apparent trend of language-impacting variation in human-divergent sequence cannot help but focus our attention on the recent evolutionary past. However, the rare variants observed in our study, which emerged even more recently to impact the language ability of our study participants, speak to the fact that the evolution of human language ability might not only be considered in the past tense, but in the present and future tenses as well.

## 5 Subjects and Methods

### 5.1 Human Subjects and IRB

All subjects in this study were minors who assented to participation under IRB# 200511767. All secondary genetic analyses were approved and carried out under the University of Iowa IRB #201406727.

### 5.2 EpiSLI Cohort

The primary discovery cohort of this study — referred to as EpiSLI — was originally recruited as a epidemiological study of language impairments in Kindergarteners in the state of Iowa. A more detailed description of the recruitment scheme can be found in the initial [64], and subsequent [64, 62] publications. In brief, 7,218 kindergarteners, sampled to be representative of Iowa’s population were screened for language impairment with a rapid, 40-item subset [64] of the TOLD-2P [48]. 26.2% of children failed this screener, and were over-sampled to 42% of a primary cohort (n = 1,929 – excluding those with reported autism spectrum disorder or intellectual disability). The entirety of this core cohort received a more complete battery of language and cognitive assessments, while teachers and parents completed scholastic and behavioral questionnaires about the children. Assessments were repeated for a portion of this core cohort at 2nd, 4th, 8th, and 10th grades. Additionally, some new recruits were added to the cohort at later ages, for a total n = 2,591 children with core language scores at a minimum of one timepoint.

### 5.3 EpiSLI Behavioral and Cognitive measures

Core language assessment scores and IQ (see Table S1) were normalized via the general scheme described in [64] to account for the over-sampling of individuals with low language ability. Core language scores were corrected for age as the residual of a linear regression model using lm() in R [51]. In cases when tests were given over two sessions, participant age was averaged across the two sessions. The age-corrected core language scores were then normalized with respect to the population using the wtd.mean() and wtd.var() functions from the Hmisc package [25] in R. Only individuals recruited as part of the epidemiological study were used to calculate the weighted means and variances used in this normalization, as later recruits did not receive the screener which determined the sampling scheme. These means and standard deviations can be found in Table S1.

#### 5.3.1 Data Imputation

Most individuals for whom whole genome sequencing was generated had scores from assessments given in 2nd grade, while a minority had only 4th grade (41/390) or kindergarten (39/390) scores. Missing scores were imputed using chained random forests and predictive mean matching as implemented the missRanger R package [42] with 5,000 trees and pmm.k = 20. See Table S1 for a summary of the final out of bag error estimate for each assessment.

#### 5.3.2 Factor Analysis

A maximum-likelihood factor analysis was carried out on the language assessment and IQ scores using the factanal() function from the stats package in R [51]. A number of factors from 1-10 were evaluated, with 7 — the largest number of factors with a nominal *χ*^2^ p-value less than 0.05 — chosen for subsequent analyses (Figure S1A). These seven factors accounted for an estimated 62.3% of the total variance in the original data (Figure S1B). For each factor, individuals were classified as scoring high, above average, below average, or low based on the weighted quartiles of each factor, accounting for the sampling scheme from the original population sample (Table S2). When binary classification of factors was required (i.e. into low-ability and typical-ability groups), a cutoff of the 33rd percentile was used.

### 5.4 Whole Genome Sequencing

#### 5.4.1 Sample Collection

DNA was collected and extracted from either blood or saliva samples. DNA concentration of all sequenced samples was quantified with with Qubit 2.0 Fluorometer (Life Technologies Corporation).

#### 5.4.2 Sequencing

390 DNA samples were sheared on a E220 Focused-ultrasonicator (Covaris) to an average size of 400 bp. Sequencing libraries were generated with a Kapa Hyper Prep kit (Kapa Biosystems) according to the manufacturer protocol. Samples were sequenced with one of two pooling schemes. Under scheme A, 8 samples were initially pooled across 8 lanes. Extra sequence was then generated ad-hoc from new pools for those samples found to have produced insufficient sequence for an average genome-wide coverage of 20X. Under scheme B samples were initially run on single lanes. Samples without an average genome-wide coverage of at least 20X were then pooled in proportion to initial mapping rate to generate additional sequence. All samples were sequenced on a HiSeq4000 (Illumina) with 150-bp Paired End chemistry.

#### 5.4.3 Post-sequencing QC

All sequence generated from all samples was analysed with Fastqc (v0.11.8), where no samples failed the run modules: Mean Sequence (4 warnings); Per-sequence Quality scores (0 warnings); Per-base sequence content (17 warnings); Per-sequence GC Content (361 warnings); Per-base N content (0 warnings); Sequence Duplication Levels (0 warnings); Over-represented Seqences (23 warnings). After genome alignment (see below), some samples were found to have significantly better coverage than others, in excess of 50x. To help control for ascertainment bias in these samples, all samples with an average coverage over 35x (based on initially mapped BAM) were randomly down-sampled, leading to the distribution of geome-wide coverage and average insert-size outlined in Supplementary Table S3

#### 5.4.4 Genome Alignment and Variant Calling

Reads were processed with bcbio (v1.1.6a-b’8be2464’) and mapped to hg19 via BWA-mem (v0.7.17). SNVs and InDels were then called with all samples in a pool with three variant callers: GATK (v4.1.2.0), FreeBayes (v1.1.0.46) and Platypus (v0.8.1.2).

Variants from each caller were filtered according to caller-specific quality metrics. For GATK, variants needed to pass all VQSR tranche thresholds. For Platypus, variants were filtered based on the goodness of fit of genotype calls, excessive region-based haplotype scores, root-mean-square mapping quality, variant quality and its ratio with read depth, low complexity sequence context, allele bias, region-based read quality, neighboring homopolymers, and strand bias. For FreeBayes, variants were filered based on a combination of allele frequency, read depth, and overall quality. The thresholds used for filtering Platypus and FreeBayes calls were the default set by bcbio.

#### 5.4.5 Assembly of High Confidence Call-set

An ensemble of all three callers was generated and used in all subsequent analyses in order to achieve improved specificity in the detection of rare variants. All variants in the ensemble callset were called by GATK as well as either Platypus OR FreeBayes. The majority of variants (87%; 25,775,508) were called by all three callers, while a minority were called by GATK and one other caller (13%; 3,937,146). Only 725,057 variants were called by platypus and FreeBayes but not GATK.

### 5.5 Variant Annotations

Sequencing variants in the high-confidence callset were annotated with the Ensembl Variant Effect Predictor (VEP v87) [43]. VEP assigns variants to various classes of coding and non-coding regions. Coding regions include genes, exons, transcripts, and CDS while non-coding regions include promoter regions, TF binding sites, motifs and more.

Additional variant annotations were also calculated using VCFanno [49] (v20190119). The deleteriousness of all single-nucleotide variants were predicted using Combined Annotation Dependent Depletion [53] (v1.4). This score combines multiple genome-wide annotations by contrasting genetic variants which persist at the population-level with simulated mutations. Allele frequency were derived from GnomAD [30] (v2.0.1), which includes the population-level allele frequencies from many whole-genome sequencing studies, but also integrates data from ExAC [31] for exonic variants. Finally, the rsIDs of variants were retrieved from dbSNP [58] (v151)

#### 5.5.1 Derivation of Technical Covariates

Starting from the initial sample size of 390, a total of 14 samples were flagged for removal from all analyses because of population stratification. Specifically, these samples did not cluster with 1,000 Genomes Europeans, or were more than 3 standard deviations away from the rest of the EpiSLI cohort based on the top 10 multidimensional scaling components calculated from SNPs found at or above a 0.05 minor allele frequency. These SNPs were extracted from the ensemble callset and merged with genotypes form 1,000 Genomes and clustered into 5 groups using PLINK (v1.90b6.6). These SNPs were also used to calculate relatedness and the top 20 common-variant principal components of the cohort using PLINK. An additional 7 samples were removed from all analyses due to cryptic relatedness with another individual in cohort (having a pi-hat greater than 0.15 as calculated by PLINK).

To control for technical effects in rare-variant analyses related to potential fluctuations in sequence quality across sequencing batches, principal components were derived using prcomp in R from a variety of sample-level metrics, namely: average coverage at variant sites, average insert size, total number of reads, mapped reads, mapped read percentage, mapped paired reads, duplicate reads, duplicate read percentage, mapped unique reads, and GC percentage. Five components were found to have nominally significant Spearman’s correlation to composite language scores (p < 0.05) were included in subsequent analyses as potential technical confounders. 19 Samples scoring more than 4 mean absolute deviations from the median of these covariates were flagged as potential technical outliers in all analyses where these confounders were included as covariates. This removal resulted in a final sample size of 350. After regressing composite language score for these technical components, three of the top 20 common genetic-variant-based principal components were also found to have nominally significant associations with the composite language score, and were included as potential confounders as well.

### 5.6 Polygenic Scores

#### 5.6.1 Genotypes

To derive a set of common-variant SNPS suitable for PGS analysis, SNPs in epiSLI found at or above a minor allele frequency of 0.05 in 1,000 Genomes were extracted from the high-confidence ensemble whole genome sequencing callset.

#### 5.6.2 Base GWASes

Polygenic scores in both ABCD and EpiSLI were calculated from neuropsychiatric GWAS summary statistics produced by the Psychiatric Genomics Consortium (PGC) for ADHD [10], Autism [24], bipolar disorder [56], schizophrenia [56] and cross disorder [38]. PGS for cognitive traits were derived from summary statistics from the Social Science Genetic Association Consortium (SSGAC) for educational attainment [37] and cognitive performance [54]. These GWAS summary statistics were downloaded directly from the respective consortia websites (SSGAC = https://www.thessgac.org/; PGC = https://www.med.unc.edu/pgc/). All summary statics were normalized to the forward strand reference and alternate alleles on human genome build hg19, with beta coefficients or odds ratios adjusted accordingly, and all ambiguous alleles removed. Due to the populations use to derive these summary statistics, only unrelated individuals of European ancestry (determined by multidimensional scaling of genotypes in PLINK [50]) were included in PGS calculations (n = 350).

#### 5.6.3 PGS Calculation and Canonical Correlation Analysis

The program PRSice v2.2.11.b [15] was used to calculate PGS for all traits. We used the linkage disequilibrium structure of 1,000 Genomes Europeans (downloaded from ftp://ftp.1000genomes.ebi.ac.uk/vol1/ftp/release/20130502/) to prune SNPs according to default parameters of PRSice because of the relatively small size of the EpiSLI cohort. PGS were calculated starting at a minimal p-value threshold of 5e-08 at increasing intervals of 1e-04. Genetic principal components and the technical covariates described above were regressed out of each PGS with glm() and z-scaled with scale() in R [51] in order to account for potential confounding influence of population substructure and technical effects. PGS scores for each trait were extracted from the PRSice output at three thresholds corresponding to approximately 5,000, 50,000, and 100,000 SNPs. The corresponding p-value thresholds and exact number of SNPs can be found in Table **??**

Regularized canonical correlation analysis was used to derived 7 correlation components between these PGS scores and the 7 language language factors via the sgcca() function from the RGCCA R package [61] with the shrinkage parameter c1 = 0.5. The direction of correlations from sgcca() are arbitrary. For consistency, the signs of loadings and fitted values were flipped so that all correlations were positive, and the sum of loadings on PGSes were positive.

### 5.7 Identification of High Impact Coding Variants

To identify putative language genes subject to high-impact variants, we utilised the VEP variant assignment and predicted effects described above in “Variant Annotations”. Variants were first filtered to those which VEP determined to have “HIGH” impact, which includes: splice variants, stop codon gains and losses, frameshift variants, and start codon losses. We required variants to be rare — found at an allele frequency less than 1% in both gnomAD v2 and the EpiSLI cohort. Given the rarity of all variants of interest, we kept only individual genotypes with a quality score greater than 20 which were supported by at least 10 reads. Finally, variants had to map to Ensembl transcripts of putative language-related genes, which we derived from literature review in [34]. We considered genes a potential hit if at least one individual with low language ability had a rare, high-impact variant in the gene, but no individuals with above average or greater ability had one.

### 5.8 Gene-level Common & Rare Variant Association

#### 5.8.1 Gene-level SKAT Test

We utilized a sequence kernel association test (SKAT) to assess the gene-level association of common and rare genetic variants with the 7 factors described above. Specifically, we used the SKAT\_CommonRare() function from SKAT package [39] in R with a fixed cutoff for common and rare variants of 0.01 We assigned variants to protein-coding genes based on VEP annotation, including variants located in introns or UTRs. To control for cohort-specific calls (i.e. sequencing artefacts) — while still allowing for potential deviation from typical populations given the cohort’s enrichment for individuals with low language ability and IQ — variants found at less than 1% in non-neuro populations of gnomAD v2 but over 5% in the EpiSLI cohort were removed, while variants found at over 1% in gnomAD v2 were allowed to occur in EpiSLI at up to 5x the gnomAD frequency. We stringently filtered individual genotypes to high-confidence calls, removing those with a genotype quality less than 20, fewer than 10 supporting reads, or more than 100 supporting reads (e.g. in the rare case of unmarked duplicates or contaminating reads). Filtered calls were considered missing, and imputed by SKAT (i.e. via Hardy-Weinberg equilibrium). Only protein-coding genes with at least 5 variants passing these filtering criteria are reported here (n = 17,782). All of the technical and population-stratification covariates described above were included in the SKAT null model.

#### 5.8.2 Replication of Common Variant Association in *NDST4* region

SNP array data was previously generated for a portion of the EpiSLI cohort, including 111 European-ancestry individuals not included in the whole-genome sequencing (WGS) cohort. Samples were genotyped on the Illumina Infinium Global Screening Array. All variants were mapped to the hg19 reference of the human genome and exported from GenomeStudio into PLINK [50] formatted files and passed through a quality control process based on [41]. Individual samples and SNPs with high global missing rates were removed in a two stage process. First, samples and SNPs with missing rates above 20% were removed, then samples and SNPs with missing rates above 5%. SNPs with MAF < 01% in the cohort were then removed, as well as those which defied Hardy-Weinberg equilibrium to a extreme degree (HWE p-value < 1^−10^). Samples with a SNP missing rate > 5% on any one autosome were then removed and the linkage disequilibrium structure of the cohort was calculated. Samples with heterozygosity rates greater 3 standard deviations from the cohort mean rate were removed, as well as samples that were outliers (3 standard deviations from the mean) on the first 10 components of multidimensional scaling of genotypes. After this QC, samples were clustered based on genotype in order to identify population substructure of the sample. The cohort was merged with the samples from 1,000 genomes phase 3 [1]. This combined set of samples was clustered into 5 groups, corresponding to the 5 super-populations found in 1,000 genomes. Finally, The top 20 principal components of each of the 5 clusters (populations) of the EspiSLI cohort were calculated separately.

To test for the association SNPs in the *NDST4* region, the genotype of the SNPs in the region for the 111 European-ancestry samples not part of the WGS were extracted from the PLINK callset and read into R [51]. A null linear model was then built, predicting factor scores based on the cohort’s top 20 genetic principal components. For each SNP, a model was built which additionally included the genotype at the evaluated SNP. The p-value of an ANOVA between the null and SNP-inclusive models are reported here.

### 5.9 Singleton Variants

#### 5.9.1 Identification of Singletons and Variant Annotations

Singleton variants — those called on only one sample in the entire cohort — were extracted from the ensemble callset. All potential singleton variants of interest had to be within 5Kb of an autosomal protein-coding gene (n = 17,434), and have an allele frequency less than 0.1 across all gnomAD V2 populations. A total of 139 annotations for these singleton variants were extracted from the database, including: reference and alternate allele length, reference and alternate allele nucleotide composition, reference and alternate allele in Chimpanzee, pathogenicity and conservation scores (i.e. CADD, FATHMM, phastCons), variants impacts from VEP (e.g. upstream, intron, missense, frameshift), predicted tissue-specific impacts [65] TisAn, 100 discrete conservation states [2] consHMM.

#### 5.9.2 Random Forest Models

##### Training Set

To establish a training set of variants, singletons assigned to genes with prior evidence for involvement in language or general cognition was curated. Putative language genes were taken from [34], and genes related to cognition were taken from the overlap of genes significantly associated with educational attainment via MAGMA [37] with genes linked to developmental disorders [6]. For training, 27 samples with an excessive burden of singletons across all genes — more than three absolute deviations above the median — were removed, leaving a maximum of 323 samples per gene. Additionally, remaining samples with an excessive burden of singletons — more than three absolute deviations above the median — in individual training-set genes were removed from the training set for those genes. The median number of samples removed per gene in this way was 14. To further minimize the impact of spurious calls, only variants where all samples in the cohort had complete calls (both alleles) were included in the training set. Finally, singletons near genes with few of these high-quality singletons (fewer than 50) were removed from the training set. The training set consisted of 53,189 singleton variants assigned to 71 gene regions.

##### Identification of Generalizable Singletons

To account for the low probability that any given singleton has any impact on cognition — or language ability specifically — the pool of training singletons were pruned to only those which generalized to samples that were held out-of-bag. To accomplish this, we utilized a leave-one-out approach at the sample level, training a random forest model to predict if a singleton belonged to an individual with a low factor score (bottom 33%) based on the annotations described above using 322 of the 323 non-outlier samples. The model was then used to predict the singletons of the 1 held-out sample, and only those variants which were predicted correctly were deemed generalizable. This process was repeated for all 323 samples and 7 factor scores, resulting in a final training sets between 27,891 and 28,485 singleton variants depending on the factor. The random forest implementation in [40] was used to train each of these models, using 100 trees and the default parameter for mtry. Stratified sampling of singletons was carried out for each tree to balance the classes (low vs typical ability) and to ensure large genes with many singletons did not have undue impact on the models.

##### Final Model Training and Risk Ratio Calculation

The singletons identified above were used to train a final random forest model for each factor. These models grew 500 trees, used the default parameter for mtry, and featured no stratified sampling. These models were then used to predict scores for the remaining singletons assigned to all other genes. The vote proportion of predictions was converted to a risk ratio using logistic regression to fit the out-of-bag vote proportion of the training set variants to their corresponding predicted class (low vs typical ability). For all final training set singletons, only the votes and predictions from the trees where that singleton was out-of-bag were carried forward.

##### Feature Importance and Association with Final Scores

For the seven final models (one per phenotype factor), the importance of each annotation was quantified as both the mean decrease in Gini index and the decrease in accuracy if the annotation was permuted. To produce a more continuous quantification of individual annotations’ associations with final risk scores — and because some annotations may quantify the same underlying biological signal — the out-of-bag scores for final training set variants were placed into 15 bins with an equal number of variants in each. Logistic regression models were then fit to each bin, predicting singletons’ membership in that bin based on each annotation. The z-scores of these models were extracted as a quantification of the annotation’s enrichment within that range of singleton risk.

#### 5.9.3 Gene-Level Enrichment

##### Gene-Level Singleton Summary Score

To identify how singleton variants associated with language ability at the gene-level, we used a weighted least squares (WLS) regression model predicting factor scores (corrected for technical covariates) from the maximal singleton risk within a gene-region. The use of WLS accounts for violations of the constant variance assumption where it occurs. Variant risk scores were taken from the out-of-bag predictions of the random forest models. Weights were computed for each singleton variant, then applied to the weighted least squares regression in R using the glm function with default parameters. Model weights were set to 1*/σ*^2^, limited between 0.1-5. *σ* was estimated in R using the lm function with default parameters to predict factor score from the singleton risk scores, then regressing out absolute values of the residuals against the singleton risk scores.

#### 5.9.4 Archetypal Analysis

To describe the functional profile of rare variants impacting each factor, archetypal analysis [16] was use to generate 20 gene-level brain-relevant archetypes, which were then correlated with the gene-level summary scores described above.

##### Gene Sets

A series of gene set lists were downloaded or processed in order to generate archetypal profiles. Summary statistics for the GWASes previously used to calculate PGS (see above) were used to generate gene-level association statistics via MAGMA [9], with default parameters and SNPs assigned to genes based on genomic location. The bulk RNA-seq expression of genes from the BrainSpan developmental transcriptome [60] were summarized to mean expression values after correction for sex and hemisphere effects. Samples from BrainSpan are derived from a variety of human brain regions and developmental time points. These were grouped according to four broad developmental periods (prenatal, early childhood, late childhood, and adulthood). Human cell-type-specific median gene expression was also downloaded from the Allen Brain Atlas [3], and expression was averaged to the cell sub-type level.

##### Optimizing Archetype Solutions

To enrich for meaningful associations between identified archetypes and gene-level singleton summary scores, archetypes were fit one thousand times with separate random seeds. Iterations which generated the most nominally significant (*p <* 0.05) archetype associations with gene-level singleton summary scores (*n* = 4) were fit and the full results are available in supplementary materials. The archetype set presented here was selected for the coherence of its relationship with respect to expression profiles and MAGMA results.

##### Archetype Enrichments

Association between singleton burden predicted to harm language ability and gene-level archetypes scores was calculated using a one-sided Pearson’s correlation (via cor.test from the R stats package). To describe functional enrichment of each archetype, we utilized the GENE2FUNC feature of FUMA [67], using the top 5% of genes for each archetype as targets and the full set of 17,434 protein coding genes as the background. The percent identity of homologous genes from chimpanzee, macaque, and mouse were extracted from the Ensembl database [71, 12] and associated with archetype scores using a two-sided Pearson’s correlation. Gene-level quantification of intolerance for synonymous, missense, and loss of function variation was downloaded from gnomAD [29] and associated with archetype scores using a two-sided Pearson’s correlation.

## Data Availability

All data produced in the present study will be made publicly available upon acceptance of the manuscript to a peer-reviewed journal.
Some data is currently available to qualified researchers via dbGaP (https://www.ncbi.nlm.nih.gov/gap/), using dbGaP Study Accession: phs002255.v1.p1

## 6 Acknowledgments

We are grateful for the contributions and of the participants of the EpiSLI cohort and their families. This work was funded by NIH grant DC014489.

## 7 Data and Code Availability

The whole genome sequencing data described here is available to qualified researches via dbGaP at: *https://www.ncbi.nlm.nih.gov/projects/gap/cgi-bin/study.cgi? study*_*i*_*d* = *phs*002255.*v*1.*p*1

The code for all analyses, figures and tables here will be available at *https://research-git.uiowa.edu/michaelson-lab-public*

## 8 Web Resources

## Notes

### Competing Interest Statement

The authors have declared no competing interest.

### Funding Statement

This study was funded by NIH grant DC014489

### Author Declarations

IRB of the University of Iowa gave ethical approval for this work.

